# Deep learning on 3D ECG geometry predicts ischemia

**DOI:** 10.1101/2025.10.29.25339046

**Authors:** Alejandro Jesús Bermejo Valdés

## Abstract

**Background:** Three-dimensional (3D) electrocardiography (ECG) is a recent methodological advance that extends the dimensionality of the standard ECG, enabling geometric descriptors that capture acute ischemia. Integrating these descriptors with deep learning (DL) may improve the discrimination between ischemic and non-ischemic states and promote the clinical translation of 3D ECG analysis.

**Methods:** ECGs from seventeen patients with acute left anterior descending (LAD) artery stenosis (>50 %) were obtained from the PTB Diagnostic ECG Database (PhysioNet). Pre- and post-catheterization recordings were analyzed in 2D and 3D (V3, V6, time) over the QRS end-T onset interval. Geometric descriptors included perimeter, curvature, three almost-curvature variants, and a newly defined torsion metric. Statistical analyses comprised univariate, bivariate, and multivariate tests (PERMANOVA), complemented by DL classification using a residual multilayer perceptron with patient-wise cross-validation, isotonic calibration, and logistic meta-blending, adopting a significance level of *α* = 0.01 (99 % confidence) to ensure inference stability given the limited sample size.

**Results:** Four descriptors changed significantly after revascularization (*P*_2*D,V* 6*t*_, *κ*_2*D,V* 6*t*_, *α*_3*D*,2_, and *τ*). Correlation analyses indicated redundancy among curvature-related metrics, whereas torsion provided independent information. PERMANOVA confirmed that torsion alone, and only metric sets including torsion, achieved significance (*p <* 0.05). The torsion-based DL model provided the best discrimination, with an area under the ROC curve of 0.76 (99 % CI, 0.57-0.94; *p* < 0.001), specificity 0.82, and a Brier score of 0.18.

**Conclusions:** The integration of torsion into a DL-based 3D ECG framework enhanced the detection of acute ischemia, increasing diagnostic specificity and improving early triage and clinical decision-making in acute cardiac care.

## 1. Introduction

The three-dimensional (3D) electrocardiographic (ECG) framework introduced by Bermejo [1] established a methodological foundation to extend classical ECG interpretation from planar voltage-time projections into a 3D spatial domain. Within this framework, cardiac electrical activity is represented as a continuous spatial trajectory across three orthogonal axes, reconstructed from conventional leads, enabling the computation of geometric descriptors that quantify the morphological complexity of voltage changes over time. Bermejo [1] introduced the *almost-curvature* metric within the polar and trivial representations of the 3D ECG framework, which appeared to detect ischemic alterations more sensitively than conventional curvature and perimeter descriptors. Almost-curvature provided enhanced responsiveness to subtle ischemic distortions not discernible in two-dimensional (2D) projections.

However, these measures alone cannot fully describe the 3D twisting behavior of the electrical deflections, an additional aspect to consider in ischemic changes. Therefore, we introduced *torsion*, a key indicator that quantifies how the 3D ECG trajectory departs from planarity [2].

Nevertheless, a critical gap remains in the ability to classify ischemic and non-ischemic states using geometric metrics derived from the 3D ECG, an approach that has not yet been explored. Addressing this challenge requires advanced analytical methodologies. In this study, we applied deep learning (DL) to evaluate the discriminative capacity of 2D and 3D ECG geometric descriptors, including perimeter, curvature, almost-curvature, and the newly defined torsion, to distinguish pre- and post-catheterization states in patients with acute left anterior descending (LAD) artery stenosis, providing a methodological basis for advanced computational and automated machine learning-based myocardial ischemia classification within the 3D ECG framework.

## 2. Methods

### 2.1. Implementation

All data processing and statistical analyses were performed in Python 3.13.3. Numerical and data operations used `numpy` and `pandas`; ECG reading and signal filtering with peak detection employed `wfdb` and `scipy.signal`. Visualization was implemented with matplotlib and seaborn. Statistical analyses used `scipy.stats` for correlation and paired tests, `scikit-learn` for standardization and performance evaluation [3], `scipy.spatial.distance` for Euclidean metrics, and `scikit-bio` for permutational multivariate analysis of variance (PERMANOVA) [4].

DL analyses were run in Python 3.12.12 using `PyTorch` and `scikit-learn`. The main model was a compact residual multilayer perceptron (MLP), while a logistic regression served as the baseline. Strict patient-wise separation was enforced using a leave-one-patient-out cross-validation (LOPOCV) strategy. Within each training fold, features were standardized on the sub-training data only, and model training employed grouped early stopping to prevent overfitting. Hyperparameters were selected through an inner cross-validation procedure without information leakage. Predictions from multiple network seeds were averaged to form an ensemble. Both neural and logistic models were probability-calibrated using isotonic regression fitted on inner validation predictions. Their calibrated outputs were then combined through a logistic meta-model trained exclusively on training data. The final decision threshold was determined using Youden’s index. Model performance was assessed using the area under the receiver operating characteristic (AUROC) curve and average precision (AP), with patient-level bootstrap 99 % confidence intervals (CI). Complementary evaluation included permutation and paired sign tests, threshold-based metrics such as accuracy, sensitivity, and specificity, and the non-thresholded Brier score. ROC and precision-recall (PR) curves were plotted with `matplotlib`.

All scripts exported results in CSV format, and the complete codebase with documentation is archived in Mendeley Data [5] to ensure reproducibility.

### 2.2. Signal acquisition and preprocessing

ECG data were obtained from the PTB Diagnostic ECG Database (PhysioNet) [6, 7]. From the 290 available subjects, only those with acute LAD stenosis >50 % were included. Each patient provided a pre-catheterization ECG recorded on day 0-1 of infarction (Precath) and at least one post-catheterization ECG (Postcath) (Table 1); records lacking catheterization dates or culprit-artery documentation were excluded.

**Table 1:**
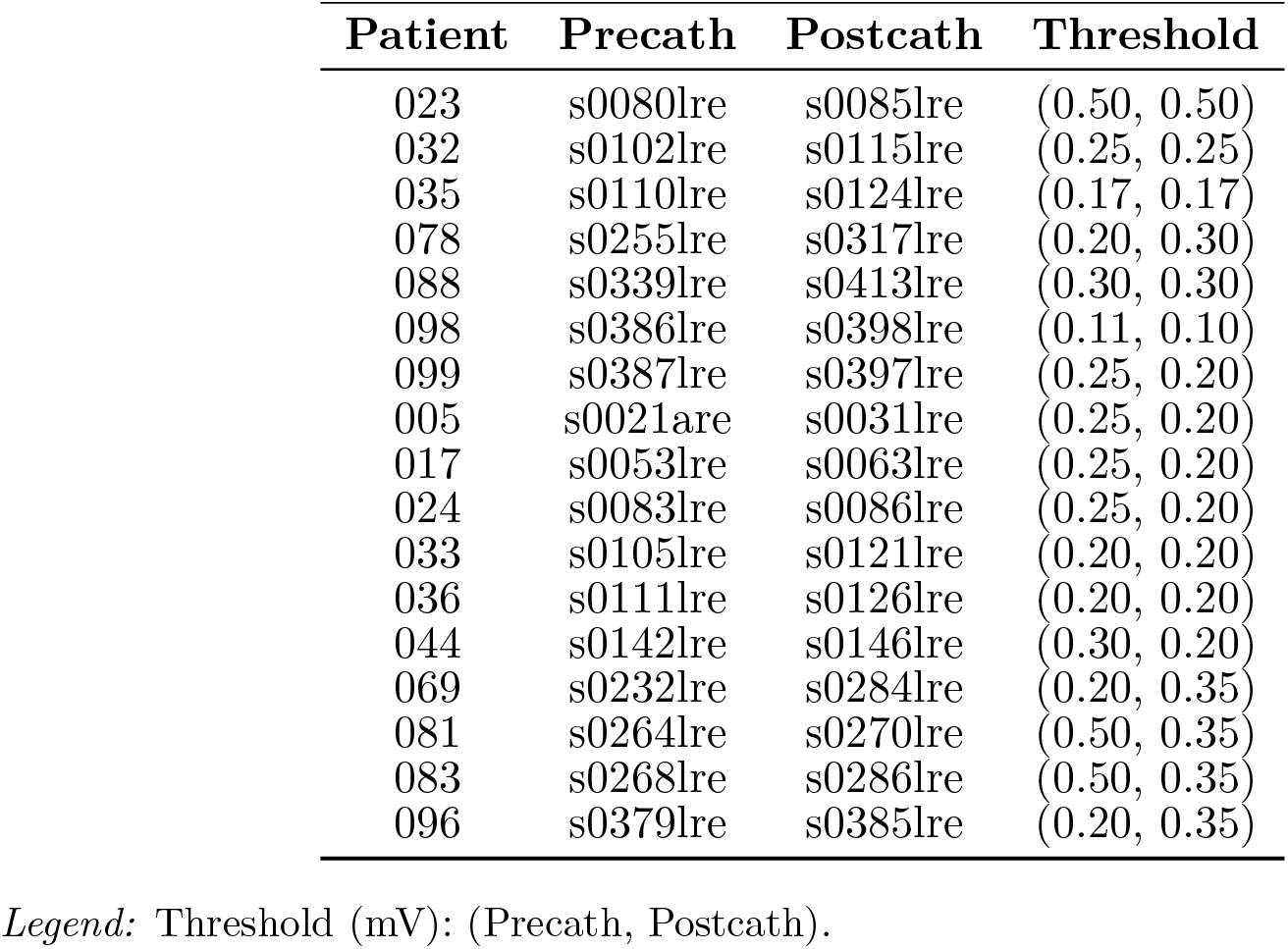
Patient and ECG record identifiers with amplitude thresholds for R-peak detection.

Signals were sampled at 1000 Hz, and a 5 s analysis window (0-5000 ms) was used for all recordings, except in patient 078 (3000-8000 ms) due to signal quality constraints. Preprocessing involved Butterworth high-pass (0.6 Hz) and low-pass (20 Hz) filtering [8].

### 2.3. Beat detection and segmentation

R-peaks were detected in lead V6 using patient-specific amplitude thresholds (Table 1), minimizing false detections from QRS fragmentation or T-wave interference. For each beat, the analysis window extended from the R-peak to one-third of the subsequent RR interval, thus adapting to heart rate variability. This interval captured the end-of-QRS to early-T phase, reducing contamination from late repolarization activity (Figures 1 and 2).

**Figure 1:**
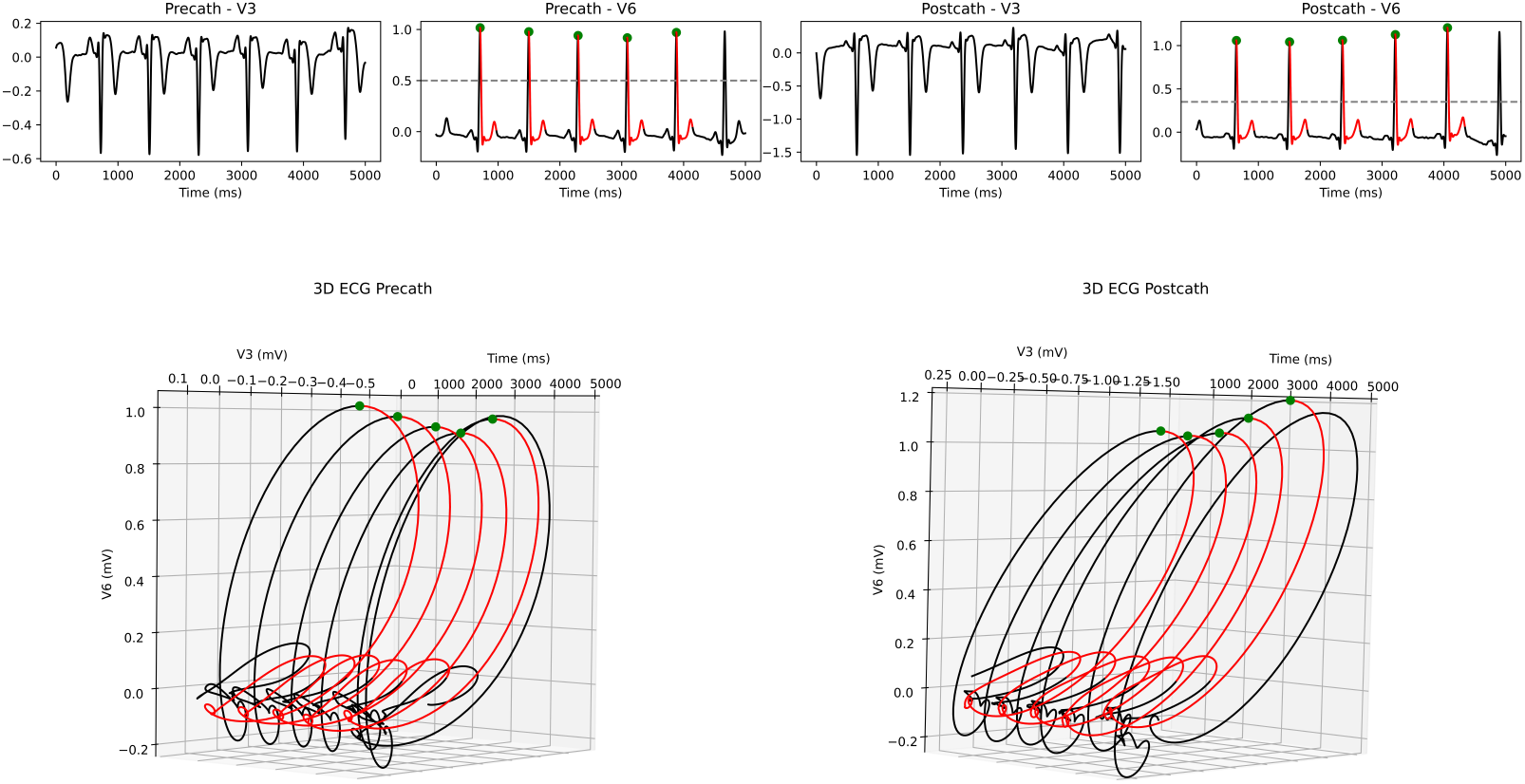
Example of 3D ECG processing in patient 081. Top: pre- and post-catheterization signals from leads V3 and V6 with R-wave detections in V6 (green markers). Bottom: reconstructed 3D ECG trajectories showing the baseline trajectory (black) and the analysis segments (red).

**Figure 2:**
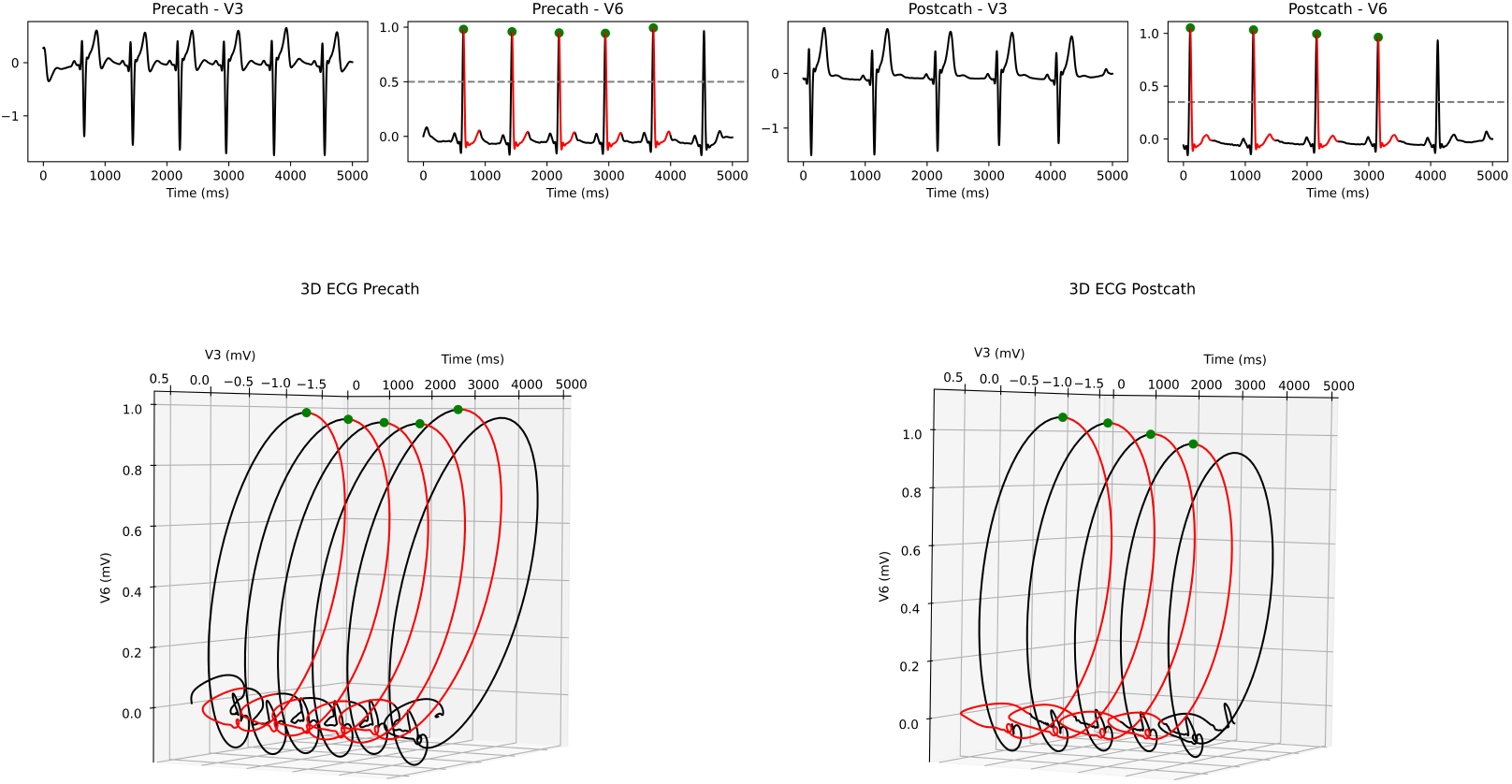
Example of 3D ECG processing in patient 083. Top: pre- and post-catheterization signals from leads V3 and V6 with R-wave detections in V6 (green markers). Bottom: reconstructed 3D ECG trajectories showing the baseline trajectory (black) and the analysis segments (red).

### 2.4. Geometric metrics

Geometric analysis was performed on both the 2D and 3D ECG trajectories reconstructed from leads V3 and V6, which are particularly informative for QRS morphology and ST-segment changes in anterolateral ischemia, the typical territory affected by LAD artery lesions [9]. Each 3D trajectory was represented parametrically as **r**(*t*) = (*x*(*t*), *y*(*t*), *z*(*t*)) = (V6(*t*), V3(*t*), *t*), following the trivial representation introduced by Bermejo [1, 10]. From the defined temporal window, multiple geometric descriptors were computed from **r**(*t*) and its planar projections. Each metric was normalized per ms by dividing by the window duration and then averaged across all valid beats within the 5 s analysis period, resulting in a single representative value per patient and descriptor. Trajectories were constructed under uniform temporal sampling Δ*t* = 1ms, so that *z*^*′*^ = 1 and *z*^*′′*^ = 0.

The perimeter *P*, curvature *κ*, and torsion *τ* were computed according to the classical formulations of differential geometry in Euclidean space [1, 2, 5], adapted to the discrete domain of sampled ECG signals. First-, second-, and third-order derivatives were estimated using discrete gradients to obtain the velocity, acceleration, and jerk vectors of the trajectory [1, 5].

The almost-curvature *α* was derived following the definition introduced by Bermejo [1], which modifies the standard *κ* by reducing the order of differentiation in one term of its numerator (Eq. 1).

The mathematical definitions of the second 3D almost-curvature variant (*α*_3*D*,2_) [1] and *τ* [2] are given by the following equations:

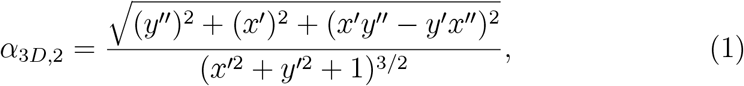

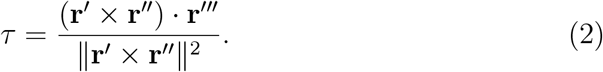

Here, the primes (*′, ′′, ′′′*) denote first-, second-, and third-order derivatives with respect to time. The cross product (*×*) defines the vector orthogonal to the plane spanned by **r***′* and **r***′′*; the dot product (·) quantifies the projection of one vector onto another; and the double bars (∥ · ∥) indicate the Euclidean norm. Under uniform temporal sampling (*z*^*′*^ = 1, *z*^*′′*^ = 0), *α*_3*D*,2_ simplifies as shown in Eq. 1.

The osculating plane at each instant is defined by the tangent and curvature vectors, **r***′*(*t*) and **r***′′*(*t*), whose normal vector is given by their cross product, **n**_osculating_ = **r***′*(*t*) *×* **r***′′*(*t*) [2]. *τ* (Eq. 2) quantifies the rate of rotation of this normal vector along the trajectory, which reflects the progressive change in orientation of the osculating plane. A torsion value of *τ* = 0 corresponds to a purely planar curve, whereas *τ* ≠ 0 indicates nonplanar 3D geometry. The sign of *τ* is orientation-dependent but not magnitude-dependent.

### 2.5. Classical statistics and multivariate analysis

For each descriptor *m*, patient-level changes were computed as the difference between pre- and post-catheterization mean values 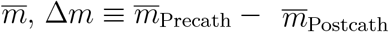.The normality of paired differences was tested with the Shapiro-Wilk test, followed by paired *t*-tests or Wilcoxon signed-rank tests as appropriate [11, 12]. Bivariate relationships between descriptors were assessed using both Pearson and Spearman correlation coefficients [13]. Overall multivariate separation between pre- and post-catheterization states was evaluated through PERMANOVA [4], applied to standardized (z-scored) descriptors using Euclidean distance matrices and 999 permutations [5]. Statistical significance was set at *α* = 0.05.

### 2.6. Deep learning framework

We implemented a leakage-free DL framework with LOPOCV, enforcing strict patient-wise separation across preprocessing, model selection, calibration, and evaluation. Within each outer training fold, features were standardized (z-score) on the sub-train partition only; validation and test data were left untouched [14]. The primary classifier was a compact residual MLP with dropout, optimized with Adam and grouped early stopping using a patient-level validation split [15]. Hyperparameters were selected via `GroupKFold` conducted exclusively within the outer training fold [3]. To reduce variance, predictions from multiple random initializations were averaged (seed ensemble). An L2-regularized logistic regression was trained on the same standardized features as a baseline model.

Probability calibration was performed by fitting isotonic regressors on inner out-of-fold (OOF) predictions derived from training data only, separately for the MLP ensemble and the logistic regression [14]. Calibrated outputs were then combined through a bagged logistic meta-model trained on training-only OOF (meta-blending) [16], mirroring the test-time prediction pipeline. For binary summaries, the operating threshold was chosen on outer-training OOF by maximizing Youden’s index [17], and subsequently applied to the held-out test patient. Reproducibility was promoted by fixing random seeds, enabling deterministic cuDNN settings when available, and exporting OOF and summary artifacts for auditability [5, 15].

All statistical estimates were computed with an error tolerance of *α* = 0.01 (99 % confidence). This stringent significance level was adopted to preserve the numerical stability of CI estimation and to ensure reliable statistical inference within the constraints of a small-sample design.

## 3. Results

### 3.1. Univariate analysis

Among all geometric descriptors, four metrics showed significant pre-versus post-catheterization differences (*p <* 0.05): *P*_2*D,V* 6*t*_, *κ*_2*D,V* 6*t*_, *α*_3*D*,2_, and *τ* (Table 2, Figure 3), confirmed by both parametric and non-parametric tests, independent of data distribution.

**Table 2:**
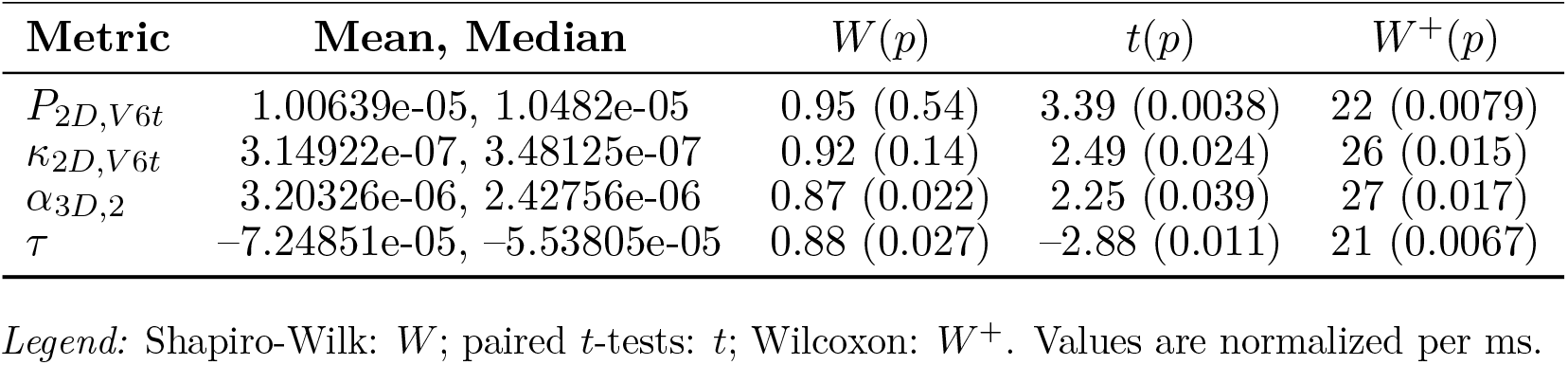
Significant pre-versus post-catheterization changes Δ*m* in geometric descriptors.

**Figure 3:**
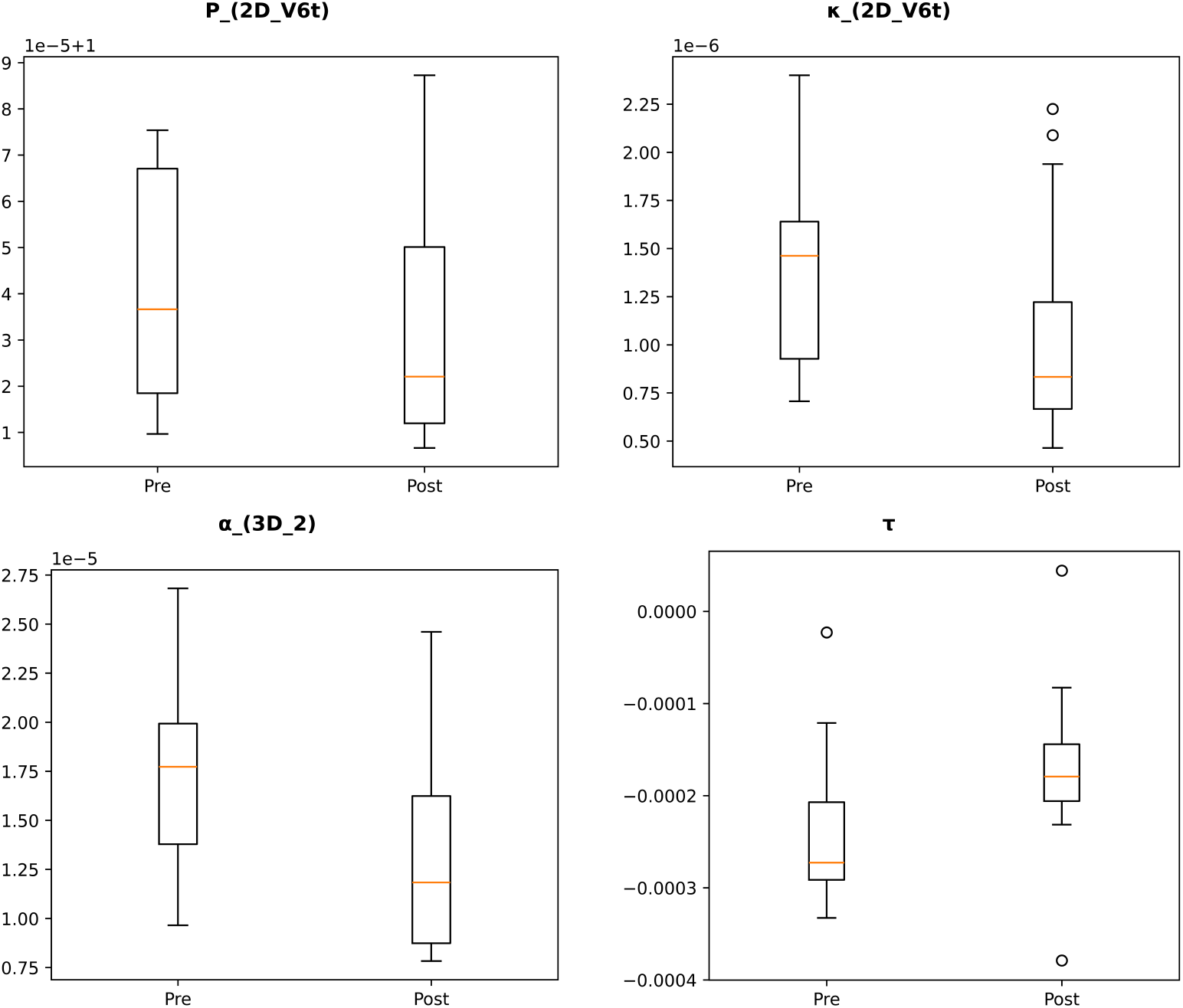
Boxplots of the four geometric descriptors that exhibited significant changes between pre- and post-catheterization states. *P*_2*D,V* 6*t*_ indicates a reduction in trajectory length within the V6-time projection. *κ*_2*D,V* 6*t*_ shows decreased curvature, reflecting smoother bending of the trajectory. *α*_3*D*,2_ displays reduced almost-curvature. *τ* remained negative in both states but shifted toward less negative values after catheterization, indicating a decrease in the magnitude of spatial twisting of the trajectory.

### 3.2. Bivariate analysis

Pairwise associations among Δ*m* revealed strong linear and monotonic relationships between *κ*-based descriptors (*κ*_2*D,V* 6*t*_ and *α*_3*D*,2_), and inverse correlations with *τ* (Tables 3–4). Concordant Pearson’s *r* and Spearman’s *ρ* values indicate that the observed associations are consistent across distributional assumptions.

**Table 3:** Pearson correlation coefficients between significant Δ*m*.

| | $P_{2D,V6t}$ | $\kappa_{2D,V6t}$ | $\alpha_{3D,2}$ | $\tau$ |
| --- | --- | --- | --- | --- |
| $P_{2D,V6t}$ | 1.0000 | 0.5536 | 0.5520 | -0.2591 |
| $\kappa_{2D,V6t}$ | 0.5536 | 1.0000 | 0.9730 | -0.6313 |
| $\alpha_{3D,2}$ | 0.5520 | 0.9730 | 1.0000 | -0.6478 |
| $\tau$ | -0.2591 | -0.6313 | -0.6478 | 1.0000 |

**Table 4:** Spearman rank correlation coefficients between significant Δ*m*.

| | $P_{2D,V6t}$ | $\kappa_{2D,V6t}$ | $\alpha_{3D,2}$ | $\tau$ |
| --- | --- | --- | --- | --- |
| $P_{2D,V6t}$ | 1.0000 | 0.6103 | 0.5564 | -0.2377 |
| $\kappa_{2D,V6t}$ | 0.6103 | 1.0000 | 0.9265 | -0.5564 |
| $\alpha_{3D,2}$ | 0.5564 | 0.9265 | 1.0000 | -0.6569 |
| $\tau$ | -0.2377 | -0.5564 | -0.6569 | 1.0000 |

### 3.3. Multivariate analysis

PERMANOVA revealed significant multivariate separation between pre- and post-catheterization states (*p <* 0.05), with the effect persisting after exclusion of *κ*_2*D,V* 6*t*_ or *α*_3*D*,2_ descriptors. Pairwise analyses showed that metric combinations including *τ* with either *P*_2*D,V* 6*t*_, *κ*_2*D,V* 6*t*_, or *α*_3*D*,2_ exhibited the strongest discriminative effects (Table 5).

**Table 5:** Global and pairwise PERMANOVA analyses across geometric descriptor sets.

| **Metric set / pair** | **pseudo-$F$** | **$p$** |
| --- | --- | --- |
| *Global models* |  |  |
| All four $\Delta m$ | 3.5262 | 0.0460 |
| Excluding $\kappa_{2D,V6t}$ | 3.6055 | 0.0320 |
| Excluding $\alpha_{3D,2}$ | 3.7002 | 0.0280 |
| *Pairwise comparisons* |  |  |
| $P_{2D,V6t}$ and $\kappa_{2D,V6t}$ | 2.3855 | 0.1210 |
| $P_{2D,V6t}$ and $\alpha_{3D,2}$ | 2.2540 | 0.1290 |
| $P_{2D,V6t}$ and $\tau$ | 3.9086 | 0.0290 |
| $\kappa_{2D,V6t}$ and $\alpha_{3D,2}$ | 3.1519 | 0.0690 |
| $\kappa_{2D,V6t}$ and $\tau$ | 4.8966 | 0.0130 |
| $\alpha_{3D,2}$ and $\tau$ | 4.7452 | 0.0230 |

### 3.4. Deep learning performance

Under LOPOCV, four 3D ECG feature sets were evaluated for ischemia versus non-ischemia classification: *τ* (SET1), *τ* + *α*_3*D*,2_ (SET2), *τ* + *κ*_2*D,V* 6*t*_ (SET3), and *τ* + *P*_2*D,V* 6*t*_ (SET4). Threshold-free performance was comparable across sets (AUROC ≈ 0.70 − 0.76, AP ≈ 0.66 − 0.75) (Table 6); SET1 achieved the highest AUROC (0.759; 99 % CI, 0.57-0.94; *p <* 0.001), while SET2 yielded the highest accuracy (0.79) and sensitivity (0.76). Specificity remained consistently high across SET1-SET3 (0.82). Calibration was stable (Brier = 0.18 − 0.21).

**Table 6:** Comparative performance of 3D ECG DL feature sets.

| SET | AUROC[99 % CI], $p$ | AP[99 % CI], $p$ | Acc / Se / Sp |
| --- | --- | --- | --- |
| SET1 | 0.760 [0.57–0.94], 0.0005 | 0.748 [0.58–0.97], 0.0009 | 0.74 / 0.65 / 0.82 |
| SET2 | 0.720 [0.52–0.93], 0.0050 | 0.710 [0.57–0.94], 0.0037 | 0.79 / 0.76 / 0.82 |
| SET3 | 0.697 [0.52–0.91], 0.0050 | 0.664 [0.54–0.92], 0.0032 | 0.74 / 0.65 / 0.82 |
| SET4 | 0.716 [0.55–0.91], 0.0010 | 0.675 [0.55–0.93], 0.0037 | 0.71 / 0.71 / 0.71 |

*Legend:* Acc = accuracy; Se = sensitivity; Sp = specificity. SET1: $\tau$ ; SET2: $\tau + \alpha_{3D,2}$ ; SET3: $\tau + \kappa_{2D,V6t}$ ; SET4: $\tau + P_{2D,V6t}$ .

According to the predefined significance level (*α* = 0.01), AUROC and AP values in all four feature sets reached statistical significance (*p <* 0.01). Among all configurations, the *τ* -only model (SET1) achieved the highest statistical significance (*p <* 0.001), supporting a very highly significant discriminative effect (Figure 4).

**Figure 4:**
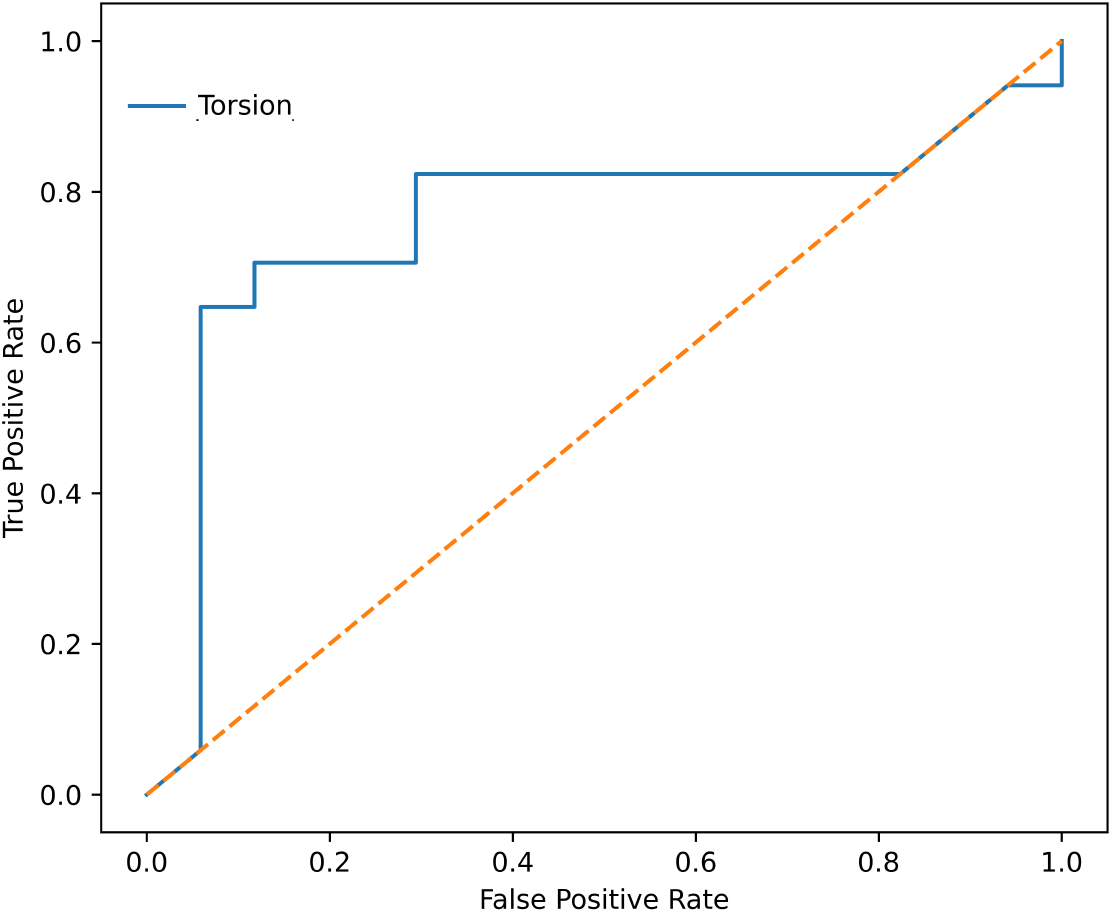
ROC curve for the DL model based on the *τ* metric, evaluated under a LOPOCV design. The x-axis shows the False Positive Rate and the y-axis the True Positive Rate, with each point representing a classification threshold. Curves approaching the upper-left corner indicate stronger discriminative performance.

## 4. Discussion

Differences in Δ*m* for *P*_2*D,V* 6*t*_, *κ*_2*D,V* 6*t*_, *α*_3*D*,2_, and *τ* indicate that acute ischemia modifies both the 2D and 3D geometries of the underlying cardiac electrical trajectory. The interdependence among descriptors reveals a layered organization of ischemic modifications: the first layer reflects the global spatial extension of the ECG deflections (*P*_2*D,V* 6*t*_); the second captures in-plane bending through *κ*-based metrics (*κ*_2*D,V* 6*t*_ and *α*_3*D*,2_); and a third layer describes the out-of-plane twisting represented by *τ*. These latter two layers appear to share specificity, with the optimal sensitivity-specificity balance and highest accuracy achieved when *τ* is combined with *α*_3*D*,2_.

Overall, the near-collinearity between *κ*_2*D,V* 6*t*_ and *α*_3*D*,2_ (*r* = 0.97, *ρ* = 0.93), their weaker associations with *τ* (|*r*| = 0.63 − 0.65, |*ρ*| = 0.56 − 0.66), and the exclusive PERMANOVA and AUROC significance of *τ* -based sets suggest that *τ* captures physiologically distinct, non-redundant information beyond *κ* and *P* metrics.

As ischemia resolves, |*τ* | decreases, indicating a reduction in the 3D twisting of the modeled 3D ECG vector and a progressive restoration of planar propagation. This attenuation of |*τ* | is consistent with the electrophysiological effects of reperfusion, including improved conduction velocity and reduced repolarization dispersion [9].

The integration of DL within the 3D ECG framework extends these observations into a predictive classification domain. Operating on precomputed geometric descriptors, the model learns nonlinear decision boundaries within the multidimensional space defined by *P*_2*D,V* 6*t*_, *κ*_2*D,V* 6*t*_, *α*_3*D*,2_, and *τ*. Through this process, the network identifies combinations of metric-derived features that best discriminate ischemic from non-ischemic states. Within this learned representation, *τ* consistently emerges as the dominant discriminant feature.

## 5. Limitations and future perspectives

This work represents an initial step toward integrating geometric descriptors and DL within a 3D ECG framework. The dataset consisted of a small and relatively homogeneous sample; however, this limitation was addressed through the use of strict LOPOCV and 99 % confidence intervals, which mitigated overfitting and ensured statistical consistency. Future research should expand the dataset size and increase sample heterogeneity, exploring additional spatial projections across other ischemic territories to further refine the detection of ischemic alterations and promote full clinical translation.

## 6. Conclusions

Reformulating the standard 2D ECG within a 3D geometric representation reveals electrophysiological information that remains hidden in conventional projections. Within this representation, *τ* emerges as a key determinant of ischemic perturbation and a physiologically meaningful marker for distinguishing acute ischemic from non-ischemic states through advanced machine learning approaches.

The attenuation of |*τ* | following reperfusion reflects the restoration of electrical coherence, reinforcing its mechanistic association with ischemic substrate distortion. Integrating *τ* and other geometric descriptors with DL represents a first methodological step toward addressing the long-standing challenge of automated ischemia detection by leveraging *τ* as a quantitative 3D marker.

This integration enhances diagnostic specificity and stability across feature sets, minimizing false-positive classifications and thereby improving triage precision in acute cardiac care. Ultimately, the 3D framework extends the diagnostic capacity of traditional ECG analysis by incorporating volumetric features of ventricular depolarization and repolarization, bridging electrophysiology with computational modeling and advancing the path toward clinically deployable 3D ECG-based decision-support systems.

Despite certain constraints inherent to the homogeneous design of this study, our findings open the door to the application of neural network models within the 3D ECG paradigm, setting the stage for future large-scale validation and clinical integration.

## Data Availability

All data analyzed in this study are openly available from the PTB Diagnostic ECG Database, hosted on PhysioNet (https://www.physionet.org/content/ptbdb/1.0.0/).
The processed datasets, Python code, and derived outputs supporting the findings of this work are available in Mendeley Data under DOI: 10.17632/md6rryc3hy.2.

